# COVID-19 Antigen Rapid Test as Screening Strategy at the Points-of-Entry: Experience in Lazio Region, Central Italy, August-October 2020

**DOI:** 10.1101/2020.11.26.20232728

**Authors:** Francesca Colavita, Francesco Vairo, Silvia Meschi, Beatrice Valli, Eleonora Lalle, Concetta Castilletti, Danilo Fusco, Giuseppe Spiga, Pierluigi Bartoletti, Simona Ursino, Maurizio Sanguinetti, Antonino Di Caro, Francesco Vaia, Giuseppe Ippolito, Maria Rosaria Capobianchi

## Abstract

COVID-19 pandemic is becoming one of the most dramatic health, social and economic global challenges in recent history. Testing is one of the main components of the public health response to contain the virus spreading. There is an urgent need to expand testing capacity and antigen rapid tests (Ag RDT) represent good candidates for point-of-care and mass surveillance testing to rapidly identify people with SARS-CoV-2 infection, counterbalancing lower sensitivity as compared to the gold standard molecular tests with timeliness of results and possibility of recurred testing. Here, we report preliminary data of the testing algorithm implemented at the points-of-entry (airports and port) in Lazio Region (Central Italy) on travelers arriving between 17^th^ of August to 15^th^ of October, 2020, using the STANDARD F COVID-19 Antigen Fluorescence ImmunoAssay. Our findings show that the probability of molecular confirmation of Ag RDT positive results is directly dependent from the semi-quantitative results of this Ag RDT, and that the molecularly confirmed samples actually harbor infectious virus. These results support the public health strategies based on early screening campaigns in settings where molecular testing is not feasible or easily accessible, using rapid and simple point of care tests, able to rapidly identify those subjects who are at highest risk of spreading SARS-CoV-2 infection.

## INTRODUCTION

As of November 15^th^, 2020, COVID-19 pandemic continues to spread worldwide accounting for a total of over 53,766,728 cases and 1,308,975 deaths, with high impact on healthcare systems and devastating global socio-economic consequences **[1]**. With COVID-19 cases accelerating towards a second wave for many Countries and a further overburden on health care systems and laboratories, there is an urgent need to expand testing capacity, in order to quickly identify as many SARS-CoV-2 positive persons as possible to control infection transmission **[2-4]**.

Nucleic acid amplification test (NAAT) is the gold standard for the diagnosis of SARS-CoV-2 infection, however it is laborious, expensive, and challenged by reagent supply shortage **[4-6]**. Antigen rapid tests (Ag RDT) can represent a good option for mass testing and for rapidly capturing individuals potentially more infectious, especially in decentralized settings, or in those scenarios where molecular testing is not feasible or easily accessible. In fact, despite the lower sensitivity, they are able to identify current infections during the most contagious phase and are faster, simpler-to-use, and less expensive than NAAT **[7-8]**. Overall, these tests may ultimately help to relieve the pressure on healthcare systems. The variability of the clinical performance is one of the limiting factors of Ag RDT **[7; 9]**, however their use may be of a greater benefit if compared to the risks associated with no testing, especially for effective and sustainable surveillance regimens **[3; 10]**. The global request for ‘test, test, test’ called attention and expectancy to these tools. Debate is ongoing amongst international public health agencies and national authorities to define the most reliable way to exploit Ag RDT **[5; 11; 12]**. As a matter of fact, how to incorporate Ag RDT in diagnostic algorithms and in public health strategies is the primary aspect to address in order to balance benefits and limitations of these tests.

Here, we describe the preliminary data from our experience in implementing Ag RDT at points-of-entry (PoE) in Rome, Italy, between 17^th^ of August to 15^th^ of October, 2020.

## METHODS

In August 2020, following the end of the national lockdown, the re-opening of borders and the intensification of citizen travels due to summer holidays, Italian authorities strengthened the border surveillance addressing all travelers returning from high incidence foreign countries, as well as from Sardinia region (Italy) **[13]**. On 17^th^ of August, in Lazio Region, on-site screening by SARS-CoV-2 Ag RDTs was implemented at the international airports in Rome (“Leonardo da Vinci International Airport”, Fiumicino, and “Ciampino–G. B. Pastine International Airport”, Ciampino), and at the port of Civitavecchia (Rome) for those ships returning from Sardinia region.

The Ag RDTs used was the STANDARD F COVID-19 Ag Fluorescence ImmunoAssay (FIA, SD Biosensor, Korea). This test detects viral nucleoprotein (N) directly from nasopharyngeal swab (NPS); the interpretation of results is performed after 30 minutes incubation using an automatic fluorescence reader (STANDARD F100, SD Biosensor, Korea) which gives a cut-off index (COI) as measure of fluorescence signal detected in relation to the presence of viral antigen; COI ≥1 is interpreted as positive for SARS-CoV-2 N antigen. The antigenic testing was voluntary and the sample collection and Ag RDT execution was performed by trained health care workers deployed at the PoE (Regional Special Unit for Community Health Care, USCAR). Based on the sensitivity and specificity of this assay, established on a preliminary validation study **[14]**, assuming 1% prevalence, positive and negative predictive values (PPV and NPV) of this test were estimated to be 23.3% (95%CI: 10.1 - 45.0) and 99.5% (95%CI: 99.4 - 99.6), respectively.

Based on this assumption, the adopted algorithm was to confirm only Ag RDT positive results. For NAAT confirmation, a second NPS was collected in UTM medium (COPAN, US) immediately after the Ag RDT results and readily sent to the Laboratory of Virology of the National Institute for Infectious Diseases “L. Spallanzani” (INMI). Different Real Time RT-PCR (RT-PCR) platforms available for the routine COVID-19 diagnosis (DiaSorin Simplexa® COVID-19 Direct; Roche Diagnostics Cobas® SARS-CoV-2; Abbott RealTime SARS-CoV-2) were used as molecular confirmatory tests. In case of negativity by Ag RDT, appropriate communication about the mandatory caution in the interpretation of the results was adopted, including recommendation of continued use of transmission prevention measures, such as mask wearing and social distancing.

Virus culture was performed on selected RT-PCR confirmed samples, using Vero E6 cell line, as previously described **[15]**.

Results of each Ag RDT were recorded at the testing site through an electronic register and, subsequently, uploaded to the Regional Surveillance Information System established by the regional health authority. Anonymized data have been extracted and analyzed using STATA 14 statistical software. Laboratory data (SARS-CoV-2 RT-PCR results) were recorded on the Laboratory Information System in use at the Laboratory of Virology of INMI; when available, COI values obtained from the Ag RDT reader at the testing site were matched with the results of molecular tests used for laboratory confirmation. For comparison between COI and viral RNA level, only samples tested with molecular platforms addressing ORF1 as viral genome target were used, in order to allow homogeneity of RT-PCR results.

Spearman correlation test was performed using GraphPad Prism version 8.00 (GraphPad 160 Software, US). Positive Predictive value (PPV) and negative predictive value (NPV) were calculated using the MedCalc statistical software (MedCalc Software Ltd, Belgium).

This work was performed within the framework of the COVID-19 outbreak response and surveillance program, and has been approved by the INMI Ethical Committee (“Comitato Etico INMI Lazzaro Spallanzani IRCCS/Comitato Etico Unico Nazionale Covid-19”; issue n. 214/20-11-2020). All data are aggregated and non-identifiable.

## RESULTS

From the 17^th^ of August to 15^th^ of October 2020, a total of 73,643 Ag RDT have been recorded in the Regional Surveillance Information System from travelers at the PoE. Of these, 72,467 resulted negative for SARS-CoV-2 N antigen, while 1,176 (1.6%) resulted positive. On the Regional Surveillance Platform, matched NAAT confirmation results were available for 941 of the 1,176 Ag RDT positive samples, resulting in 381 (40.5%) RT-PCR positive confirmation. The proportion of Ag RDT confirmed results by NAAT was well within the 95%CI of the expected proportion, on the basis of the PPV calculated for 1% prevalence (see the Methods section).

The analysis was performed on a sub-set (n=603) of samples from subjects tested positive by Ag RDT with available COI values **(Fig. 1)**.

**Figure 1.**
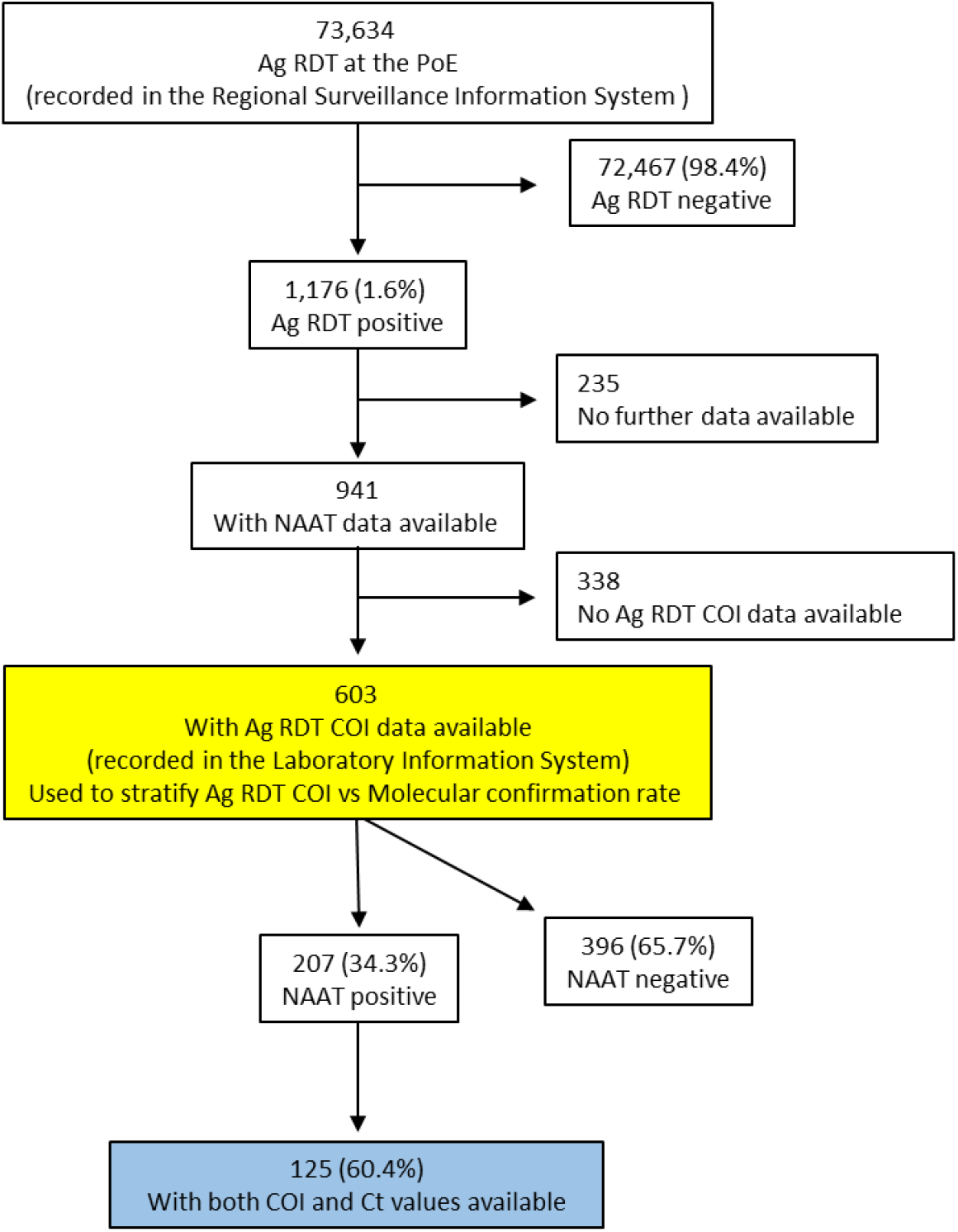
Flow-chart of the analysis performed on the records from Ag RDT testing at PoE and NAAT laboratory confirmation. The yellow and blue boxes correspond to the data analysed in Table 1 and Fig. 2, respectively.

The COI range of this sub-set was 1.01-87.7. Of these samples, 207 (34.3%) were confirmed positive by SARS-COV-2 RT-PCR. For 125 samples both Ct ORF1 and COI values were available. The median Ct value was 19.8 (range 11.1-34), corresponding to high viral loads in NPS which are potentially more infectious **[5]**. Viral culture on VERO E6 cells was attempted on 10 of these NPS resulted positive by Ag RDT and confirmed by SARS-COV-2 RT-PCR (Ct range: 11.4-19.1); replication-competent virus was recovered from all of these samples, supporting the potential for SARS-CoV-2 transmission. In addition, a significant correlation (r = −0.6; p<0.0001) was observed between the Ct values, that are surrogate markers of viral load (lower Ct value corresponds to high viral load) and COI values resulted from the Ag RDT, which represent the extent of the antigen detection **(Fig. 2)**.

**Figure 2.**
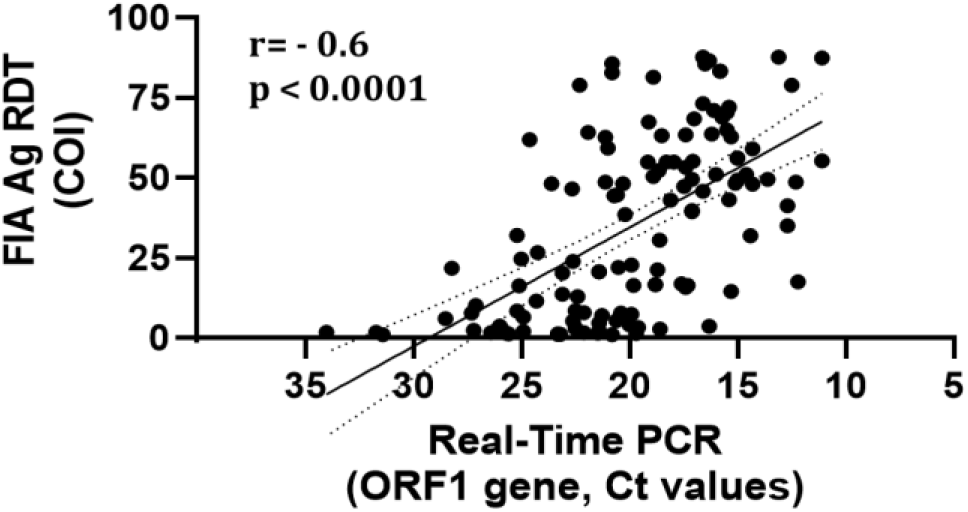
Correlation between RT-PCR Ct values and COI obtained on confirmed SARS-CoV-2 positive samples with available information for both parameters (n= 125, as in blue box of Fig. 1).

In fact, the median COI value of the Ag RDT not confirmed by SARS-CoV-2 RT-PCR (n=397) was very low (1.4, range: 1.0-15.0). We then stratified the RT-PCR result confirmation rate according to the COI obtained by Ag RDT. As shown in **Table 1**, the percentage of confirmed Ag RDT results was strongly dependent on the COI value, ranging from 81.6% when the COI threshold was set at 3 to 100% when the COI threshold was set at 20. More in details, positive Ag RDTs with COI≥10 were confirmed in 98.7% of cases with only 1.3% of Ag RDT false positive results, while lower cut-off showed higher percentage of Ag RDT false positivity.

**Table 1.** Percentage of confirmed Ag RDT positive results by SARS-CoV-2 RT-PCR according to the COI values (n=603, as in yellow box of Fig. 1).

| **COI** | **RT-PCR positive / N**<br>**(% TP)** | **% FP**<br>**by Ag RDT** |
| --- | --- | --- |
| ≥1 | 206/603 (34.2%) | 65.8% |
| ≥3 | 186/228 (81.6%) | 18.4% |
| ≥5 | 175/188 (93.1%) | 6.9% |
| ≥8 | 159/165 (96.4%) | 3.6% |
| ≥10 | 152/154 (98.7%) | 1.3% |
| ≥15 | 138/139 (99.3%) | 0.7% |
| ≥20 | 127/127 (100%) | 0.0% |

Abbreviation: COI, cut-off index; TP, true positive; FP, false positive

## DISCUSSION

In this study, we evaluated the results of the SARS-CoV-2 testing strategy at PoE in Lazio Region using Ag RDT in order to reduce the risk of importing cases and limit new chains of transmission. The test was voluntary and the tool implemented was the STANDARD F COVID-19 Ag FIA (SD Biosensor, Korea), characterized by 30 minutes to results and the use of automatic user-friendly reader which guarantees objective interpretation of the results. More than 70,000 travelers arriving to airports and port of Lazio Region were screened by Ag RDT between 17^th^ of August and 15^th^ of October 2020, and a small proportion (1.6%) resulted positive for SARS-CoV-2 antigen. The proportion of samples confirmed with RT-PCR was 40.5%, with almost 60% resulting false positive as expected in low prevalence settings **[5; 12]**.

As for other Ag RDT, a previous study showed that STANDARD F COVID-19 Ag FIA is highly specific (98.4%, 95%CI: 96.0% to 99.6%) for SARS–CoV-2 Ag detection in NPS, and highly sensitive (95.2%, 95% CI: 76.2% to 99.9%) for those samples with Ct values lower than 25, which are harbored by patients with high viral load, more likely expected to be able to transmit the infection **[14; 16; 17]**. In our experience, high viral load values (median Ct value=19.9) were observed in samples confirmed positive by NAAT. More importantly, our study shows that clinical samples resulted positive by Ag RDT and confirmed by NAAT harbor infectious virus. To our knowledge, this is the first report showing a linear correlation between viral loads in NPS (measured as Ct values) and extent of viral antigen detection (measured as COI values). As a consequence, the proportion of Ag RDT samples confirmed by a molecular test was extremely high (98.7%) in those samples with COI ≥10, reaching 100% with COI ≥20. This information may be of help for a reliable diagnostic interpretation of this specific tool and may be useful for a potential reassessment of the diagnostic algorithms of Ag RDT confirmation by NAAT. In fact, on the basis of the proportion of cases confirmed by NAAT according to their COI value, a threshold (e.g. COI≥10) may be established to exempt molecular testing when the expected confirmation rate exceeds 95%. Following this principle, it is possible that specific thresholds may be evaluated also for other semi-quantitative Ag RDT, prompting not only a consequent earlier implementation of public health measures, but also a potential advantage for the NAAT laboratory workload and demand in terms of staff effort and reagents supply, ultimately helping to relieve the pressure on healthcare systems **[3-4; 11; 16]**.

A limitation of the present study is that Ag RDTs resulted negative at PoE did not undergo RT-PCR testing, so it was not possible to estimate the proportion of PCR-positive samples missed by Ag RDT. However, in our opinion, though Ag RDTs are substantially less sensitive than NAAT **[8]**, they are worth to be integrated into COVID-19 outbreak management programs, as they may contribute to the prompt isolation of highly infectious cases who would otherwise have been lost, highlighting the role of PoE testing as a pillar of outbreak control **[11; 18]**. In fact, the use of Ag RDT may be particularly advantageous to early minimize the risk of the virus spreading, especially when and where there is no immediate access to RT-PCR testing, or where this cannot be feasible, as for mass and frequent testing or in certain field settings. In addition, Ag RDT use could also represent a suitable screening tool for prompt cluster investigation and in specific cohorts such as asymptomatic contacts of COVID-19 confirmed cases, pauci-symptomatic patients with no epidemiological link and travelers with no symptoms **[11]**. Of course, public-awareness campaigns must also communicate that a negative Ag RDT does not necessarily implies a clean bill of health, and continued social distancing and mask wearing needs to be recommended. In addition, negative Ag RDT results in symptomatic patients should not be considered definitive, and molecular tests are required for a more reliable diagnostic assessment. Finally, it is necessary to balance clinical diagnostic performances of the tests, testing sustainability (i.e. cost, staff demand, technology and infrastructures, time to results), public health implications, and socio-economic consequences, which can also derive from delay in diagnostic response **[3; 19]**.

## Data Availability

Data available unpon request.

## Acknowledgments

We acknowledge the *Local Health Authorities of Lazio Region*; the *Port and Airport Health Office (USMAF) of Lazio Region*; the *Covid-19 INMI Study Group* (Maria Alessandra Abbonizio, Chiara Agrati, Alessandro Agresta, Fabrizio Albarello, Gioia Amadei, Alessandra Amendola, Mario Antonini, Raffaella Barbaro, Barbara Bartolini, Martina Benigni, Nazario Bevilacqua, Licia Bordi, Veronica Bordoni, Marta Branca, Paolo Campioni, Maria Rosaria Capobianchi, Cinzia Caporale, Ilaria Caravella, Fabrizio Carletti, Concetta Castilletti, Roberta Chiappini, Carmine Ciaralli, Francesca Colavita, Angela Corpolongo, Massimo Cristofaro, Salvatore Curiale, Alessandra D’Abramo, Cristina Dantimi, Alessia De Angelis, Giada De Angelis, Gabriella De Carli, Rachele Di Lorenzo, Federica Di Stefano, Federica Ferraro, Lorena Fiorentini, Andrea Frustaci, Paola Gallì, Gabriele Garotto, Maria Letizia Giancola, Filippo Giansante, Emanuela Giombini, Maria Cristina Greci, Giuseppe Ippolito, Eleonora Lalle, Simone Lanini, Daniele Lapa, Luciana Lepore, Andrea Lucia, Franco Lufrani, Manuela Macchione, Alessandra Marani, Luisa Marchioni, Andrea Mariano, Maria Cristina Marini, Micaela Maritti, Giulia Matusali, Silvia Meschi, Francesco Messina, Chiara Montaldo, Silvia Murachelli, Emanuele Nicastri, Roberto Noto, Claudia Palazzolo, Emanuele Pallini, Virgilio Passeri, Federico Pelliccioni, Antonella Petrecchia, Ada Petrone, Nicola Petrosillo, Elisa Pianura, Maria Pisciotta, Silvia Pittalis, Costanza Proietti, Vincenzo Puro, Gabriele Rinonapoli, Martina Rueca, Alessandra Sacchi, Francesco Sanasi, Carmen Santagata, Silvana Scarcia, Vincenzo Schininà, Paola Scognamiglio, Laura Scorzolini, Martina Spaziante, Giulia Stazi, Francesco Vaia, Francesco Vairo, Maria Beatrice Valli); *INMI Covid-19 laboratory team* (Abbate Isabella, Agrati Chiara, Aleo Loredana, Alonzi Tonino, Amendola Alessandra, Apollonio Claudia, Arduini Nicolina, Bartolini Barbara, Berno Giulia, Biancone Silvia, Biava Mirella, Bibbò Angela, Bordi Licia, Brega Carla, Canali Marco, Cannas Angela, Capobianchi Maria Rosaria, Carletti Fabrizio, Carrara Stefania, Casetti Rita, Castilletti Concetta, Chiappini Roberta, Ciafrone Lucia, Cimini Eleonora, Coen Sabrina, Colavita Francesca, Condello Rossella, Coppola Antonio, D’arezzo Silvia, Di Caro Antonino, Di Filippo Stefania, Di Giuli Chiara, Fabeni Lavinia, Felici Luisa, Ferraioli Valeria, Forbici Federica, Garbuglia Anna Rosa, Giombini Emanuela, Gori Caterina, Graziano Silvia, Gruber Cesare Ernesto Maria, Khouri Daniele, Lalle Eleonora, Lapa Daniele, Leone Barbara, Marchetti Federica, Marsella Patrizia, Massimino Chiara, Matusali Giulia, Mazzarelli Antonio, Meschi Silvia, Messina Francesco, Minosse Claudia, Montaldo Claudia, Neri Stefania, Nisii Carla, Petrivelli Elisabetta, Petroni Fabrizio, Petruccioli Elisa, Pisciotta Marina, Pizzi Daniele, Prota Gianluca, Raparelli Fabrizio, Rozera Gabriella, Rueca Martina, Sabatini Rossella, Sarti Silvia, Sberna Giuseppe, Sciamanna Roberta, Selleri Marina, Selvaggi Carla, Sias Catia, Stellitano Chiara, Toffoletti Antonietta, Truffa Silvia, Turchi Federica, Valli Maria Beatrice, Venditti Carolina, Vescovo Tiziana, Vincenti Donatella, Vulcano Antonella, Zambelli Emma).

